# Antibody responses following COVID-19 vaccination and breakthrough infections in naïve and convalescent individuals suggests imprinting to the ancestral strain of SARS-CoV-2

**DOI:** 10.1101/2025.05.19.25327882

**Authors:** Siddhartha Mahanty, Emily M. Eriksson, Peta Edler, Francesca Mordant, Nicholas Kiernan-Walker, David J Price, Sabine Braat, Eamon Conway, Vanessa Bryant, Honghua Ding, Leo Yi Yang Lee, Louise Randall, Ramin Mazhari, COVID PROFILE consortium, Ivo Mueller, Kanta Subbarao

## Abstract

The binding and neutralising activity of SARS-CoV-2 antibodies are important correlates of protection of current COVID-19 vaccines. SARS-CoV-2 exposure status and COVID-19 vaccine types can influence these responses and the breadth of cross-reactivity to variants. In this longitudinal cohort study, we used SARS-CoV-2-specific multiplex Luminex^®^ antibody assays and live virus neutralisation of ancestral (VIC01/2020), Delta and Omicron (BA1, BA2 and BA5) SARS-CoV-2 variants to compare antigen-specific binding and neutralising antibody (nAb) responses to primary vaccination (two doses) of adenovirus vectored (AdVV) or mRNA vaccines followed by a booster dose of mRNA vaccine in convalescent (n=51) and infection-naïve individuals (n=47). In a subset of individuals, we performed additional analysis of antibody responses following breakthrough infection.

We found that titres of anti-SARS-CoV-2 nAb following primary vaccination (2 doses) with AdVV vaccine were significantly lower than those following mRNA vaccine, irrespective of prior SARS-CoV-2 infection status. However, an mRNA vaccine booster dose resulted in equivalent binding and nAb titres to the ancestral virus in all individuals, irrespective of primary vaccine type. Notably, vaccinated infection-naïve, but not convalescent individuals required the third dose of vaccine (mRNA) to induce nAbs to Omicron subvariants BA1, BA2 and BA5, though titres against the variants were lower than those against the ancestral strain. Importantly, breakthrough infection with Omicron strains induced higher nAb titre rises against the ancestral strain than against Omicron variants consistent with imprinting of the immunologic response and recall of pre-existing immunity to the ancestral strain.

## Introduction

The COVID-19 pandemic has presented varying challenges across the globe, with Australia experiencing a unique trajectory. With public health measures, notably, stringent border closures, mandatory quarantine for arriving travellers and social distancing, Australia maintained a low case count during the initial phases of the pandemic^1–4^. This period, preceding the emergence of the Omicron variants in late 2021, allowed for widespread vaccination efforts, leveraging mRNA and adenovirus-vectored vaccines, achieving a remarkable vaccination rate of >90% among adults. Consequently, with the re-opening of the borders and emergence of Omicron strains, Australia witnessed a low incidence of severe COVID-19 cases within a largely immunized population^5^. This distinctive scenario provided an exceptional opportunity to investigate immune responses to SARS-CoV-2 vaccination without the confounding influence of background immunity from prior infections. In addition, it facilitated a comparative analysis of antibody responses against emerging variants among vaccinated individuals with and without previous exposure to the virus.

Early reports have highlighted the emergence of anti-SARS-CoV-2 antibodies shortly after infection, with subsequent dynamics characterized by a rapid initial decline followed by a more gradual decay in titres^6–9^. Studies investigating vaccine-induced antibody kinetics, particularly in response to mRNA, protein, and vector-based COVID-19 vaccines, have demonstrated robust responses, albeit with considerable variation in peak levels and decay rates across vaccine types ^10–13^. Notably, individuals with prior COVID-19 infection exhibit significantly elevated antibody levels post-vaccination, with a slower decline over time^14^.

Neutralizing antibodies (nAb) generated by vaccination with the spike protein from the ancestral virus prevent virus entry into host cells efficiently, but levels of cross-reactivity with SARS-CoV-2 variants vary, irrespective of whether the nAbs are generated by vaccination or by natural infection^15–17^. However, the majority of serological studies, have been conducted in settings with high community transmission of the virus^18^, making it difficult to evaluate neutralising activity generated by COVID vaccines over extended infection- free periods that were characteristic of low-transmission environments like Australia^3,19,20^.

In this cohort study, we explored SARS-CoV-2 antibody responses among COVID-19-naïve and convalescent individuals in Australia following vaccination with BNT162b2 (mRNA) or ChAdOX1 (AdVV) vaccines. Our investigation includes evaluation of antibody levels and nAb against ancestral, Delta and Omicron (BA1, BA2 and BA5) variants, following primary immunization, booster doses given months after the primary vaccines, and around breakthrough infections with variant viruses. Our findings support recent reports of the dominance of neutralizing activity against the ancestral strain post-vaccination^21–24^, and yield insights into the development of heterologous (AdVV primary vaccine and mRNA booster) versus homologous (all vaccines were mRNA type) immunity against SARS-CoV-2 variants following vaccination and breakthrough infections.

Our results illuminate the complex interplay between vaccination, prior infection, and emerging variants, shedding light on the dynamics of antibody responses crucial for informing ongoing pandemic response strategies.

## Materials and Methods

### Study subjects

This study is an immunological sub-study from two study cohorts - DISCOVER-HCP-Vaccine cohort (Peter Doherty Institute for Infection and Immunity, Melbourne, Australia, Melbourne Australia) (N=95) and COVID PROFILE cohort^25^ (The Walter and Eliza Hall Institute, Melbourne Australia) (N=171). From the two cohorts, a total of 126 sera and plasma samples were collected from 57 participants in the DISCOVER-HCP-Vaccine cohort and from 69 participants in the COVID PROFILE cohort. Study cohort demographics are detailed in supplementary data (**Table S1a**). At study entry participants were categorised into infection-naïve controls (Naive; n=72) or COVID-recovered (Convalescent; n=54). COVID-19 infection status was determined based on available results of PCR tests for SARS-CoV-2 RNA in nasal/oral swabs and further verified by SARS-CoV-2-specific antibody levels at baseline (pre-vaccination). Ninety eight participants (51 Naïve and 47 Convalescent individuals) who had received 2 doses of COVID-19 vaccines at the start of the current immunological sub-study and also had pre-vaccination samples collected and another 4 (Naïve) prior to the booster vaccine were included in the analysis of responses to the primary vaccines and booster dose. The analysis of pre- and post-infection samples included data from 61 participants (36 Naïve and 25 Convalescent participants). Demographics for this subpopulation is summarised in **Table S1b**. These studies were approved by the Walter and Eliza Hall Institute (#20/08), Melbourne Health (RMH69108), and Royal Melbourne Hospital Human Research Ethics Committee (HREC #63096/MH-2020). All individuals provided written informed consent to participate in this study, in accordance with the Declaration of Helsinki.

### Vaccination, sample collection and breakthrough infection

All participants received either two doses of BTN162b2 (mRNA), or ChAdOX1 (AdVV) under an Australian government-supported vaccination program. When a third (booster) dose was recommended, a subset of participants (n=115) received one dose of mRNA vaccine. Ninety-eight participants entered the study prior to receiving the first vaccine dose, 4 entered before the booster vaccine and 24 entered after the primary vaccine and had a breakthrough infection. Depending on the timepoint relative to vaccination when participants were enrolled in the study, serum or plasma samples were collected prior to vaccination (Pre-Vax) as a baseline sample, and 2-4 weeks after the second dose of vaccine (Post-2^nd^ Vax) and/or 2-4 weeks after the third vaccine dose (Post-3^rd^ Vax). The last sample collected prior to the third vaccine dose served as pre-3^rd^ Vax. A number of participants (N=61) had a breakthrough infection during follow-up. The last sample collected before the breakthrough diagnosis served as pre-infection sample and post-infection samples were collected 2-4 weeks post-diagnosis of the infection (**Fig. S1**).

Sample flow for SARS-CoV-2-specific binding antibody and nAb titre analyses is summarised in Supplementary **Figures S2 and S3** respectively.

### SARS-CoV-2-specific antibody multiplex assay

Plasma antibody levels specific for SARS-CoV-2 antigens S1, S2, receptor- binding domain (RBD), Spike and nucleoprotein (NP), based on sequences from the ancestral strain, Delta RBD, Omicron BA1 and BA2 RBD were measured using a multiplex serological assay employing the Luminex platform as previously described^26^. Antibody levels to seasonal coronavirus antigens (NL63 NP, OC43 Spike, 229E S1 and HKU1 Spike), Influenza A antigen (H1N1 haemagglutinin) and tetanus toxoid were also measured in the assay. For each individual, total IgG, IgM and IgA levels were measured for each ancestral strain-derived antigen and total IgG levels were measured for variant antigens. For standardisation between plates data were normalised using an algorithm which adjusted for plate-to-plate variation based on standard curves.

### Micro-neutralisation assay (MNT)

SARS-CoV-2 isolates, including CoV/Australia/VIC01/2020 (the ancestral strain)^27^ were passaged in Vero cells and Omicron variants (specific strains BA.1, BA.2 and BA.5) were passaged on TMPRSS-expressing Vero cells and stored at −80°C. Serum samples were heat-inactivated at 56°C for 30 minutes. Serial dilutions of plasma, ranging from 1:20 to 1:10240, were prepared before the addition of 100 TCID50 of the respective SARS-CoV-2 variant in MEM/0.5% BSA. The mixtures were incubated at room temperature for 1 hour. Residual virus infectivity in the serum/virus mixtures was assessed in quadruplicate wells of Vero cells or TMPRSS-expressing Vero cells, as appropriate. The cells were incubated in serum-free media containing 1 μg/ml of TPCK trypsin at 37°C and 5% CO2 and viral cytopathic effect was evaluated on day 5. The nAb titre was calculated using the Reed–Muench method as previously described^28,29^.

### Statistical Analysis

The sample sizes used in our analyses were constrained by the number of individuals with available data in each of the two sources of participants, the DISCOVER-HCP and the COVID Profile studies. Antibody measures that were below the limit of detection were assumed to be the limit of detection (i.e., a titre of 10 in the microneutralisation assay), such that fold-rise measures were conservative, and the corresponding estimates provide a lower-bound on the true fold change.

A linear mixed-effects regression model was used to calculate the geometric means at each time point and the fold-change of the geometric means between two specified time points, (with corresponding 95% confidence intervals (CI)). The outcome was the log antibody levels (RBD binding or micro neutralisation assay titre (MNT)) and the models included fixed effects for timepoint, vaccination type (AdVV or mRNA), pre-study infection status (naïve or convalescent) and for MNT outcomes, and the virus variant (ancestral, Delta, BA1, BA2 or BA5). Repeated measures of individuals were accounted for via a random effect (intercept) for each participant. To obtain estimates by the vaccine type received and pre-infection status (and COVID-19 variant for MNT outcomes), all two-, three- and four-way interaction terms were also included in the model.

The emmeans function (emmeans package in R; ^30^) was used to estimate the geometric mean at each time point from the model as a marginal mean effect. The margins command (margins package in R^31^) was used to estimate the fold-change in antibody levels between two time-points as the marginal effect. Corresponding two-sided 95% confidence intervals (95% CI) and p-values for pre-infection status, vaccine type, and antibody type combination from the linear mixed-effects model are reported. 95% CI will be quoted herein as (95% CI [lower limit, upper limit]). Statistical significance was assigned to p-values ≤ 0.05. No adjustment for multiple testing was applied to the confidence intervals or p-values given that the outcomes were not powered for.

Correlation of binding antibody to neutralising antibody levels was calculated using Spearman rank correlation, with 95% CIs calculated via z-transformation.

## Results

### Demographic distribution across study cohort groups

Overall, there was a higher number (68-72%) of females in all study groups, except the Convalescent AdVV group, where only 40% were female (**Table S1a**). The median age for participants receiving the AdVV vaccine (52.0 years for Naïve and 59.0 years for Convalescent) was higher compared to participants who received the Pfizer primary vaccine (39.0 years for Naïve and 46.0 years for Convalescent). This is as expected, as Australians over 50 years of age were eligible for only the AdVV vaccine as part of the initial vaccine rollout in Australia^32^. On average, around 40% of participants who received an AdVV primary vaccine had a breakthrough infection, as opposed to 52-57% of participants who received the mRNA vaccine. However, the overall percentage of participants who experienced a breakthrough infection was similar for the Naïve group compared to Convalescent group (50% compared to 46%; **Table S1b**).

### Robust SARS-CoV-2-specific antibody levels after two doses of COVID-19 vaccination in both previously uninfected and convalescent individuals

Vaccine-induced antibody responses have been the subject of several studies which have provided valuable insights into the immune response to COVID-19 vaccination^33–36^. To determine if the vaccine-induced immune responses in our low-transmission study cohort align with previous observations, where robust antibody responses have been described after two COVID-19 vaccine doses, we measured IgG, IgM and IgA antibody levels to several Spike protein-derived SARS-CoV-2 antigens including the receptor binding domain (RBD; **Fig. 1A**), S1, S2, Spike trimer and the nucleoprotein (**Fig. S4**) in previously SARS-CoV-2 uninfected (naïve) individuals and in individuals who had recovered from SARS-CoV-2 infection (convalescent). Antibody levels for all isotypes assessed prior to vaccination (Pre-Vax) in convalescent individuals were tested a median of 233 days (range 153 to 422) days after initial diagnosis and median of 27 days (range 8 to 73) after a second dose of COVID-19 vaccination (Post-2^nd^ vax). The naive and convalescent groups were further stratified based on which COVID-19 vaccine was received in the primary vaccination, AdVV or mRNA. Compared to pre-vaccination levels, we found that two doses of vaccine resulted in an increase in antibody levels to ancestral RBD-specific IgG antibody levels in all individuals (**Fig. 1A**, top panel). However, the fold-change in RBD-specific IgG levels from pre- vaccination to post-2^nd^ vaccine was greater in naïve participants that received AdVV (geometric mean (GM) 63.3 fold, 95% CI [38.5, 104.3], p<0.001) or mRNA (GM: 150.3 fold, 95% CI [100.7, 224.3], p<0.001) compared to convalescent participants who received an equivalent primary vaccination of either AdVV (GM 5.3 fold, 95% CI [3.4, 8.4], p<0.001) or mRNA (GM 6.9 fold, 95% CI [4.3, 11.0], p<0.001; **Fig 1 and Table S2a**). This higher fold-change meant that despite the absence of prior antigen-exposure before vaccination, IgG antibody levels to SARS-CoV-2 RBD post-2^nd^ vaccine in naïve participants were not substantially different from corresponding levels in convalescent participants, vaccinated after recovery from infection, for either AdVV (infection-naïve participants: GM of the median fluorescence intensity (MFI) 11155, 95% CI [7520, 16548] and convalescent GM MFI 17988, 95% CI [12550, 25782]) or mRNA vaccine (naïve participants: GM MFI 16705 95% CI [12169, 22930] and convalescent: 23603, 95% CI [16341, 34095], **Table S2a**). Generally, ancestral-RBD-specific IgM antibody levels (**Fig. S5, top panel**) increased after two vaccine doses for convalescent participants of either AdVV (GM fold-change over pre-vaccination level 2.4, 95% CI [1.7, 3.6], p<0.001) or mRNA vaccine (GM fold change 1.9, 95% CI [1.3, 2.9], p<0.001 (**Table S2a**). In contrast, no evidence of change in the IgM antibody levels was observed after two doses of vaccine in naïve participants that received AdVV (0.9 GM fold-change 95% CI [0.6, 1.4], p=0.652) or mRNA vaccine (1.2 GM fold-change 95% CI [0.8, 1.6], p=0.369) (**Fig. S5 top panel, Table S2a**).

**Figure 1.**
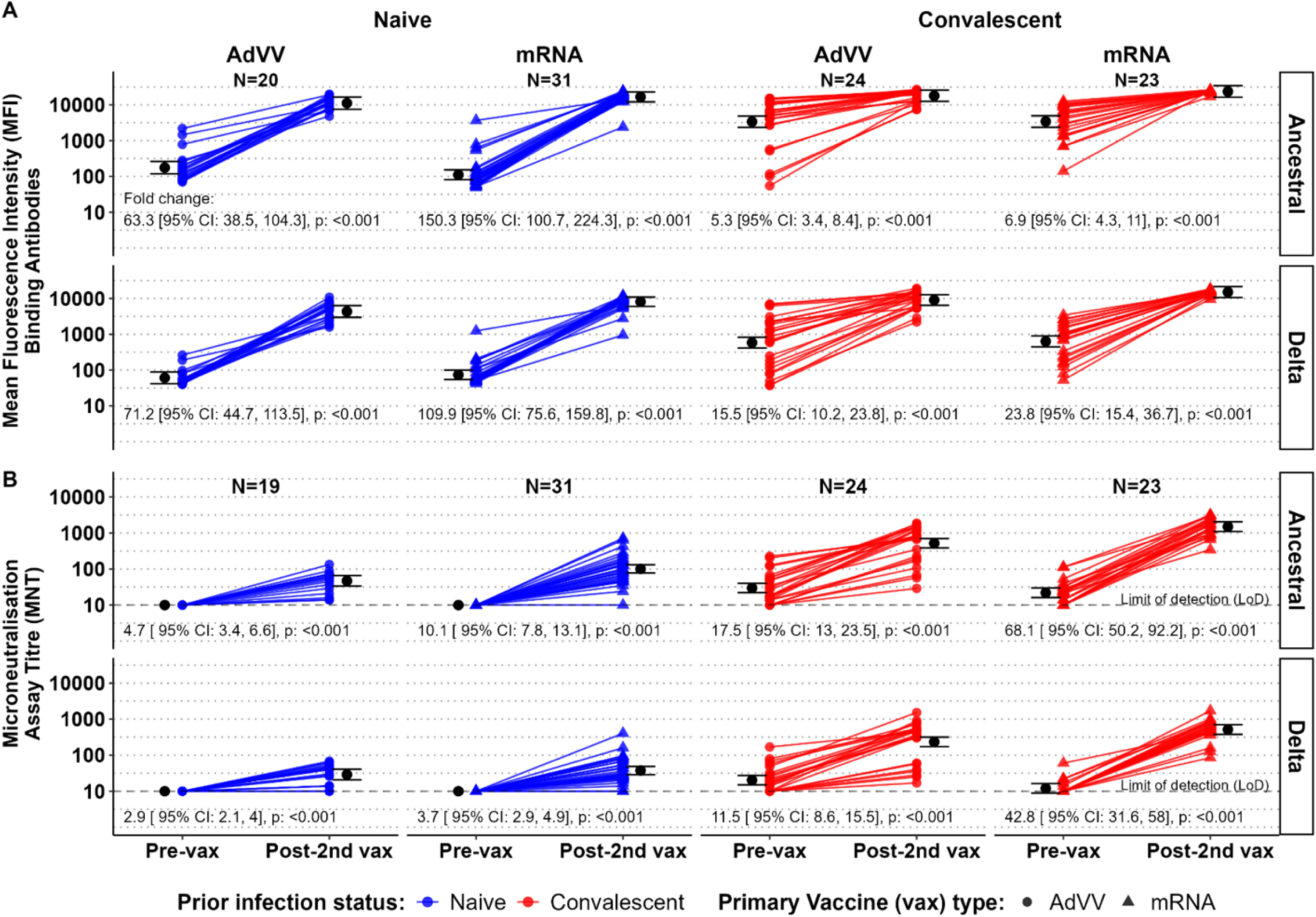
High levels of anti-RBD IgG binding and neutralising antibodies in response to two doses of COVID-19 vaccines. Anti-RBD IgG binding antibodies (expressed as median fluorescence intensity [MFI] values) (**A**) to ancestral (top panel) and Delta RBD (second panel) and neutralising antibody titres (**B**) to the ancestral strain (top panel) and Delta variant (bottom panel) at pre-vaccination (pre-vax) and post-2^nd^ vaccination (post-2^nd^ vax) time points for each individual. Participants are stratified based on vaccine type (AdVV shown as filled circles and mRNA, as filled triangles) and prior infection status (blue for naïve participants and red for convalescent participants). The geometric mean and 95% confidence interval (CI) for each time point, within each sub-group are shown in black. The geometric fold change from pre-vax to post-2^nd^ vax antibody levels, with 95% CI and p- values are shown under each plot.

While vaccination generally resulted in an increase in ancestral-RBD-specific IgA levels after two doses of vaccine (**Fig. S5 bottom panel**), naïve (6.2 GM fold-change 95% CI [4.5, 8.4]) and convalescent individuals (2.4 GM fold- change 95% CI [1.6, 3.4]), who received mRNA vaccine, had greater fold- changes in ancestral RBD-specific IgA antibodies, compared to participants receiving AdVV vaccine (Naïve: 1.6 GM fold-change 95% CI [1.1, 2.4] and Convalescent: 1.4 GM fold-change 95% CI [1.0, 2.1]) (**Fig. S5 bottom panel, Table S2a**).

Collectively these findings established that antigen-specific IgG binding antibodies are induced by COVID-19 vaccination irrespective of vaccine type, but antibody responses following two doses of mRNA vaccine were higher than following two doses of AdVV vaccine. Notably, fold-changes in antibody levels between pre-and post- vaccination timepoints were most prominent in the SARS-CoV-2 naïve population, leading to antibody levels post-second vaccine being comparable between naïve and convalescent participants (**Table S2b**). Furthermore, increases in levels of antigen-specific IgA and IgM binding antibodies were observed to a lesser extent than for IgG and were not statistically significant for IgM.

### Vaccination induces neutralising antibodies against the wildtype ancestral strain

While total antigen-specific antibody levels are a good measure of an overall humoral response elicited by COVID-19 vaccines, neutralising antibodies (nAb) have been reported to be a correlate of protective immunity to SARS- CoV-2^10^. Therefore, in addition to binding antibody levels, we also determined neutralising activity against live wildtype (ancestral) virus, in sera from each individual before vaccination (pre-vax) and after two doses of COVID-19 vaccine (post 2^nd^ vax; **Fig. 1B top panel**). NAbs were detected after vaccination in both convalescent and naïve individuals, but the vaccine response was higher among the convalescent individuals. Average (geometric mean) titres of nAbs against the ancestral strain increased 4.7 fold (95% CI [3.4, 6.6], p<0.001) in the naïve group and 17.5 fold (95% CI [13, 23.5], p<0.001) in the convalescent group following two doses of AdVV vaccine. Naïve individuals who were vaccinated with two doses of mRNA vaccine had an average increase of 10.1 fold in GMT (95% CI [7.8, 13.1], p<0.001) from pre-vaccination levels (**Fig. 1B top panel, Table S3**). The corresponding GM fold-increase in the convalescent group was 68.1 (95% CI [50.2, 92.2], p<0.001). Of note, the fold-change in nAb titres from pre-vaccination to post-second vaccine was 3.7 times (95% CI [2.4, 5.8]) greater in convalescent participants than naïve participants for AdVV vaccine recipients and 6.7 times (95% CI [4.5, 10.1]) for mRNA recipients. Furthermore, after a second vaccine dose, the average (geometric mean) nAb titres in participants who received mRNA vaccines were 2.1 times (95% CI [1.4, 3.3]) higher for naïve and 2.9 times (95% CI [1.9, 4.4]) higher for convalescent individuals compared to participants who received AdVV vaccine (**Table S3**).

While high antigen-specific antibody levels do not always equate to high nAb titres, we found that 2-5 weeks after 2 doses of COVID-19 vaccines total RBD-specific IgG levels was strongly correlated with nAb titres in naïve (r=0.65, 95% CI [0.44, 0.79]) and convalescent individuals (r=0.85, 95% CI [0.76, 0.91]; **Fig. S6**).

### Two doses of COVID-19 vaccines induce lower neutralising activity to the Delta variant than to the ancestral strain of SARS-CoV-2 despite high binding antibody levels

By December 2021, the Delta variant of SARS-CoV-2 had been circulating in Australia for ∼6 months and the Omicron variant replaced it as the dominant circulating variant^5,37^. At this time, approximately 87% of eligible Australians had received 2 doses of a COVID-19 vaccine^38^. All of the participants in our study had received two doses of COVID-19 vaccines. Analysis of antibodies to the variants in our study cohort revealed that, like antibody-binding to the wildtype-derived RBD, there was a robust rise in total IgG antibody levels that bound to the Delta-derived RBD after 2 doses of COVID-19 vaccines in both vaccine groups (**Fig. 1A**, **bottom panel**). However, the GM fold-change in levels from pre-vaccination to post-2^nd^ vaccine naïve participants was 4.6 times greater than in convalescent participants for both the AdVV (95% CI [2.4, 8.6]) and mRNA vaccine groups (95% CI [2.6, 8.2]) (**Table S4**) which reflects higher average pre-vaccination levels due to prior exposure in the convalescent group. Consequently, despite the greater fold increase, the average MFI antibody level post-2^nd^ vaccine for naïve participants was, as expected, about half of the average level for convalescent participants (**Table S4**).

When comparing whether there were differences in binding capacity to Delta RBD post-2^nd^ vaccine between individuals receiving mRNA or AdVV vaccine, we found that average Delta-RBD-binding antibodies were 1.9 (95% CI [1.1, 3.1]) times higher in naïve mRNA recipients compared to naïve AdVV vaccine recipients and 1.7 (95% CI [1.0, 2.7]) times higher in convalescent participants who received mRNA compared to the corresponding AdVV vaccine recipients (**Table S4**).

As with the neutralising titres against the ancestral strain, increases in cross- reactive neutralising titres to the Delta variant were also observed after two doses of vaccine (**Fig. 1B**, **bottom panel**). The average fold-change in GMT against the Delta variant was greater in convalescent participants compared to naïve individuals for both AdVV (4.0 fold, 95% CI [2.5, 6.2] and mRNA vaccine recipients (11.4 fold, 95% CI [7.7, 17.1]). However, the GMT of vaccine-induced neutralising activity against the wildtype variant was 1.6 – 2.9 times higher compared to the Delta variant (**Table S5**)

### Primary vaccination and a third vaccine dose result in equivalent IgG levels to Omicron subvariants BA1 and BA2

The Australian government recommended a third dose of COVID-19 vaccine to boost immunity in the population preceding the Omicron variant infection wave in Australia. Irrespective of the primary vaccine received (mRNA or AdVV), participants in our study received a third dose of COVID-19 vaccine using the mRNA formulation containing the S-protein sequence from the ancestral strain. We evaluated antibody responses to Omicron subvariants in our study participants 2-5 weeks after their third vaccine dose.

Here we assessed the levels of IgG Abs binding to the RBD protein antigen derived from either BA1 or BA2 subvariants after primary vaccination (post-2^nd^vax), prior to vaccine dose three (pre-3^rd^ vax) and after the third vaccine dose (post-3^rd^ vax) in uninfected individuals that were enrolled as SARS-CoV-2 naïve and infected individuals who were convalescent from an infection with the ancestral strain (**Fig. 2A**).

**Figure 2.**
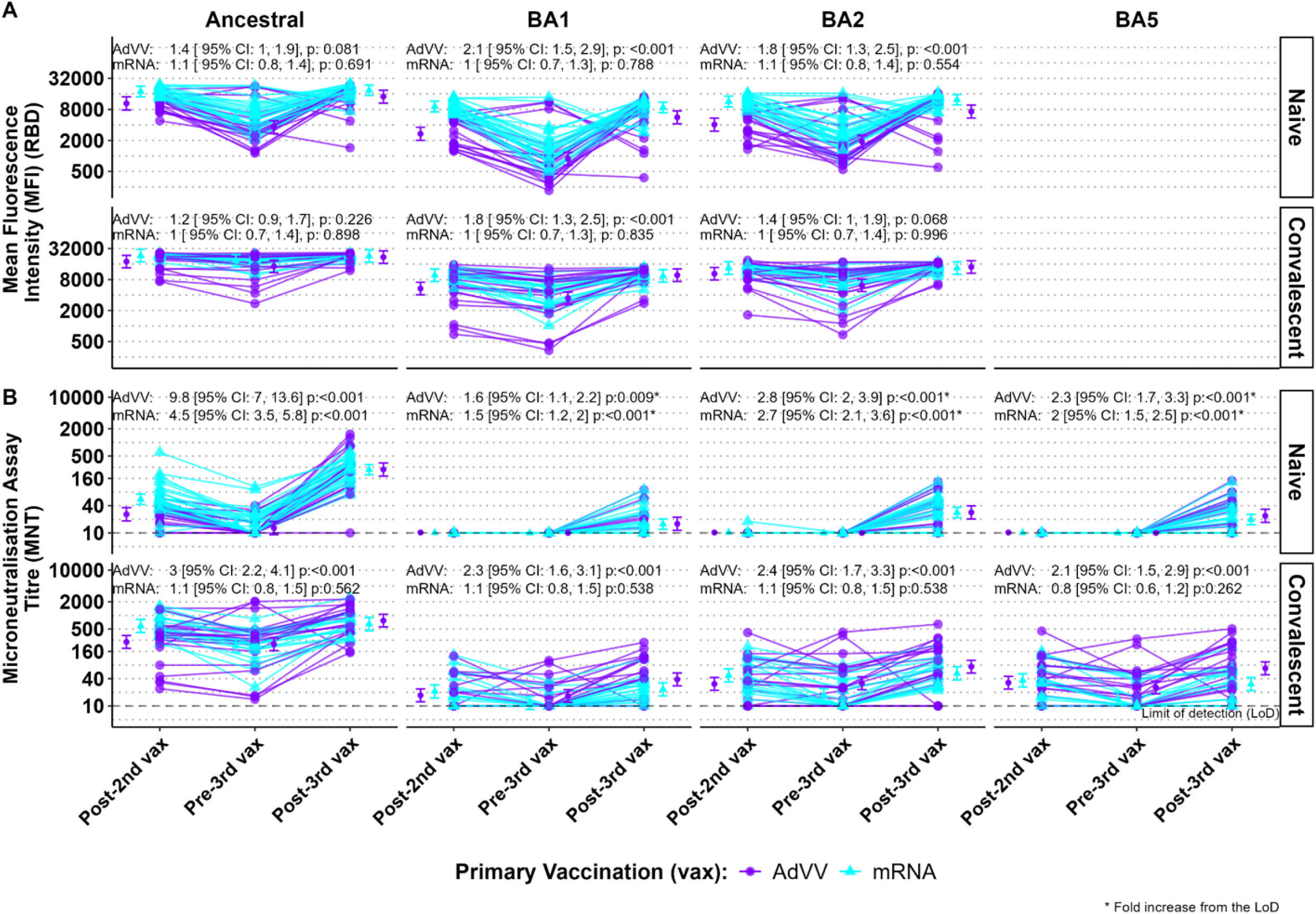
Boosting of anti-RBD IgG binding antibodies mean fluorescence intensity (MFI) levels and neutralising antibody titres after three doses of COVID-19 vaccines in naïve and convalescent individuals. Anti-RBD IgG binding antibodies (**A**) and neutralising antibody titres (**B**) at post-2^nd^ vaccination (post-2^nd^ vax), pre-3^rd^ vaccination (pre-3^rd^ vax) and post-3^rd^ vaccination (post-3^rd^ vax) time points for the ancestral strain and Omicron sub-variants (BA1, BA2 and BA5), for each individual, based on their prior infection status (naïve and convalescent). Results are shown separately for each primary vaccination type (purple for AdVV and aqua for mRNA). The geometric mean and 95% confidence interval (CI) for each time point, within each sub-group are shown by the dot and error bars. The fold- change, with 95% CI and p-value, from post-2nd vaccine to post-3^rd^ vaccine are indicated for each primary vaccine type above the plot.

We found that the levels of IgG binding to RBD from both subvariants were higher overall in naïve individuals who received two doses (primary vaccination) of the ancestral strain-derived mRNA vaccine (BA1 MFI GM 9102, 95% CI [7159, 11573] and BA2 (MFI GM 11344, 95% CI [8922, 14423]) compared to recipients of AdVV (BA1 MFI GM 2690 95% CI [2006, 3606] and BA2 (MFI GM 4067, 95% CI [3034, 5453]; **Table 1**). However, the vaccine subgroups (AdVV vs. mRNA recipients) did not differ within the convalescent cohort (**Table 1**) apart for binding to BA1 RBD (AdVV MFI GM 5346 95% CI [4044, 7068 and mRNA MFI GM 9713 95% CI [7347, 12841]). After primary vaccination at the timepoint between dose 2 and 3 (pre-3^rd^ vax), the overall antigen-specific antibody levels had declined. However, average BA2 binding antibody levels were generally higher than BA1 binding antibody levels at this timepoint (**Table 1**).

**Table 1.** Changes in levels of antigen-specific IgG binding antibodies to SARS-CoV-2 RBD proteins from Ancestral, BA1 and BA2 variant viruses following 2 and 3 doses of COVID-19 vaccines.

| Primary vaccination + booster type | Original infection | N | SARS-CoV-2 sub-variant | Geometric Mean MFI<sup>†</sup><br>[95% Confidence interval] |  | Fold change [95% CI]<sup>††</sup><br>From pre-3rd to post-3rd vaccine |
| --- | --- | --- | --- | --- | --- | --- |
|  |  |  |  | Post-2nd Vaccine | Post-3rd Vaccine |  |
| AdVV + mRNA | Naive | 17 | Ancestral | 10438<br>[7786, 13993] | 14111<br>[10592, 18798] | 1.4 [1, 1.9], p= 0.081 |
| AdVV + mRNA | Naive | 17 | BA1 | 2690<br>[2006, 3606] | 5629<br>[4226, 7500] | 2.1 [1.5, 2.9], p<0.001 |
| AdVV + mRNA | Naive | 17 | BA2 | 4067<br>[3034, 5453] | 7278<br>[5463, 9696] | 1.8 [1.3, 2.5], p<0.001 |
| AdVV + mRNA | Convalescent | 19 | Ancestral | 17837<br>[13492, 23581] | 21790<br>[16482, 28807] | 1.2 [0.9, 1.7], p=0.226 |
| AdVV + mRNA | Convalescent | 19 | BA1 | 5346 [4044, 7068] | 9736<br>[7364, 12871] | 1.8 [1.3, 2.5], p<0.001 |
| AdVV + mRNA | Convalescent | 19 | BA2 | 10391<br>[7860, 13738] | 14049<br>[10627, 18573] | 1.4 [1.0, 1.9], p=0.068 |
| mRNA + mRNA | Naive | 25 | Ancestral | 17794<br>[13995, 22625] | 18816<br>[14950, 23681] | 1.1 [0.8, 1.4], p=0.691 |
| mRNA + mRNA | Naive | 25 | BA1 | 9102<br>[7159, 11573] | 8765<br>[6964, 11031] | 1.0 [0.7, 1.3], p=0.788 |
| mRNA + mRNA | Naive | 25 | BA2 | 11344<br>[8922, 14423] | 12329<br>[9796, 15517] | 1.1 [0.8, 1.4], p=0.554 |
| mRNA + mRNA | Convalescent | 19 | Ancestral | 23361<br>[17670, 30883] | 22869<br>[17298, 30233] | 1.0 [0.7, 1.4], p=0.898 |
| mRNA + mRNA | Convalescent | 19 | BA1 | 9713<br>[7347, 12841] | 9385<br>[7099, 12407] | 1.0 [0.7, 1.3], p=0.835 |
| mRNA + mRNA | Convalescent | 19 | BA2 | 13399<br>[10135, 17714] | 13410<br>[10144, 17729] | 1.0 [0.7, 1.4], p=0.996 |

<sup>†</sup> MFI: Median fluorescence intensity; CI: confidence interval
<sup>††</sup> Using a regression model (detailed in Statistical analysis section), p-value for difference from 1.0, signifying no change, in MFI values

Interestingly, in homologous vaccine recipients (mRNA for both primary vaccine series and third dose), for both naïve and convalescent groups, the average binding antibody levels for both Omicron sub-variants BA1 and BA2, were similar after two doses compared to average levels after three vaccine doses (**Fig 2A and Table 1**). However, in recipients of heterologous vaccine (AdVV for primary vaccine series and mRNA for the third dose), binding IgG levels were approximately 2-fold higher after the third dose compared to levels after two doses of vaccine in naïve participants for BA1 (2.1 GM fold change 95% CI [1.5, 2.9]) and BA2 (1.8 GM fold change 95% CI [1.3, 2.5]; **Table 1**). For the corresponding convalescent subgroups, the fold-change between the two vaccine events was slightly lower for BA1 (1.8 fold change 95% CI [1.3, 2.5]; **Fig 2A and Table 1**).

Binding antibody levels to both subvariants after three mRNA doses (homologous vaccination) in naïve participants were similar to those seen in convalescent participants. In participants receiving heterologous vaccination, binding antibodies to both subvariants were lower in naïve participants compared to convalescent (**Table 1**). These data indicate that, for participants receiving AdVV vaccine, hybrid immunity against the ancestral strain is associated with higher binding antibody levels to Omicron subvariants compared to immunity generated by vaccine alone (naive).

### Neutralising antibody titres against SARS-CoV-2 variants are significantly boosted by a third vaccine dose in naïve individuals

Since our study participants were vaccinated with the ancestral strain-derived S protein, we investigated whether primary vaccination followed by a booster (third) dose generated neutralising activity against emerging new Omicron subvariants BA1, BA2 and BA5. Sera collected after primary vaccination (post-2^nd^ vax), pre-vaccine dose 3 (pre-3^rd^ vax) and post third vaccine dose (post 3^rd^ vax) from naïve and convalescent individuals were tested in a MNT assay utilising the ancestral strain or one of the Omicron variants as target (**Fig. 2B**). After primary vaccination in naïve individuals there was a complete absence of detectable neutralising activity to all Omicron subvariants tested (**Fig. 2B**). However, the third vaccine dose generated detectable neutralising activity against Omicron subvariants in this cohort (**Fig. 2B and Table 2**). In convalescent participants, who had detectable nAb levels after 2 doses, we found that 3 doses of mRNA vaccine (homologous vaccination; post-3^rd^ vax) resulted in similar neutralising activity against all variants as was observed after 2 vaccine doses (post-2^nd^ vax), indicating a restoration of antibody levels without significant boosting by the third dose. In contrast, heterologous vaccination in convalescent participants induced 2-3 times higher nAb titres against all variants after the third vaccine dose. (post-3^rd^ vax, **Fig. 2B and Table 2**) compared to titres after 2 doses (post-2^nd^ vax).

**Table 2.** Changes in neutralising antibody titres against SARS-CoV-2 Ancestral, BA1, BA2 and BA5 variant viruses following 2 and 3 doses of COVID-19 vaccines.

| Primary vaccination + booster type | Original infection status | Number of participants | COVID-19 Sub-variant | GMT [95% CI]<sup>†</sup> |  | Fold changes in GMT [95% CI]<sup>†</sup> |  |
| --- | --- | --- | --- | --- | --- | --- | --- |
|  |  |  |  | Post-2nd Vaccine | Post-3rd Vaccine | From vaccine 2 to post-vaccine 3 | Between variant and ancestral strains |
| AdVV + mRNA | Naive | 19 | Ancestral | 25.8<br>[18.4, 36.2] | 252.5<br>[180.8, 352.7] | 9.8<br>[7.0, 13.6], p<0.001 | Reference |
| AdVV + mRNA | Naive | 19 | BA1 | 10* | 15.9<br>[11.4, 22.2] | 1.6<br>[1.1, 2.2], p=0.009 | 0.06<br>[0.05, 0.09], p<0.001<sup>††</sup> |
| AdVV + mRNA | Naive | 19 | BA2 | 10* | 28.4<br>[20.3, 39.7] | 2.8 [2.0, 3.9],<br>p<0.001 | 0.11<br>[0.08, 0.16], p<0.001 |
| AdVV + mRNA | Naive | 19 | BA5 | 10* | 23.9<br>[17.1, 33.4] | 2.3<br>[1.7, 3.3], p<0.001 | 0.09 [0.07, 0.13],<br>p<0.001 |
| mRNA + mRNA | Naive | 31 | Ancestral | 55.8<br>[42.9, 72.6] | 249.7<br>[192.3, 324.4] | 4.5<br>[3.5, 5.8], p<0.001 | Reference |
| mRNA + mRNA | Naive | 31 | BA1 | 10* | 15.6<br>[12.0, 20.31] | 1.5<br>[1.2, 2.0], p<0.001 | 0.06<br>[0.05, 0.08], p<0.001 |
| mRNA + mRNA | Naive | 31 | BA2 | 10* | 28.3<br>[21.8, 36.8] | 2.7<br>[2.1, 3.6], p<0.001 | 0.11<br>[0.09, 0.15], p<0.001 |
| mRNA + mRNA | Naive | 31 | BA5 | 10* | 19.8<br>[15.2, 25.7] | 2.0<br>[1.5, 2.5], p<0.001 | 0.08<br>[0.06, 0.1], p<0.001 |
| AdVV + mRNA | Convalescent | 20 | Ancestral | 260<br>[187.7, 360.1] | 770<br>[556, 1066.3] | 3.0<br>[2.2, 4.1], p<0.001 | Reference |
| AdVV + mRNA | Convalescent | 20 | BA1 | 17.1<br>[12.4, 23.7] | 38.6<br>[27.9, 53.5] | 2.3<br>[1.6, 3.1], p<0.001 | 0.05<br>[0.04, 0.07], p<0.001 |
| AdVV + mRNA | Convalescent | 20 | BA2 | 30.7<br>[22.2, 42.6] | 73.7<br>[53.3, 102.1] | 2.4<br>[1.7, 3.3], p<0.001 | 0.10<br>[0.07, 0.13], p<0.001 |
| AdVV + mRNA | Convalescent | 20 | BA5 | 32.6<br>[23.6, 45.2] | 67.8<br>[49.0, 93.9] | 2.1<br>[1.5, 2.9], p<0.001 | 0.09<br>[0.06, 0.12], p<0.001 |
| mRNA + mRNA | Convalescent | 19 | Ancestral | 586.1<br>[419.6, 818.5] | 645.9<br>[462.5, 902] | 1.1<br>[0.8, 1.5], p=0.562 | Reference |
| mRNA + mRNA | Convalescent | 19 | BA1 | 21<br>[15.0, 29.3] | 23.3<br>[16.7, 32.5] | 1 .1<br>[0.8, 1.5], p=0.538 | 0.04<br>[0.03, 0.05], p<0.001 |
| mRNA + mRNA | Convalescent | 19 | BA2 | 47.3<br>[33.9, 66.11] | 52.5<br>[37.6, 73.31] | 1 .1<br>[0.8, 1.5], p=0.538 | 0.08<br>[0.06, 0.11], p<0.001 |
| mRNA + mRNA | Convalescent | 19 | BA5 | 36.7<br>[26.3, 51.31] | 30.4<br>[21 .8, 42.5] | 0.8<br>[0.6, 1.2], p=0.262 | 0.05<br>[0.03, 0.07], p<0.001<sup>..</sup> |

\*Limit of detection in microneutralization assay
<sup>†</sup> GMT: Geometric mean neutralisation titre; CI: confidence interval
<sup>††</sup> Using a regression model (detailed in the Statistical analysis section), p-value for difference from 1.0, signifying no change, in GMT values

After the third vaccine dose, nAb titres were similar for heterologous and homologous vaccination for the BA2 subvariant (**Table 2**) for both naïve and convalescent groups. In contrast, for subvariants BA1 and BA5, in convalescent participants, heterologous vaccine recipients had higher nAb titres (GMT for BA1 38.6, 95% CI [27.9, 53.5]; for BA5 67.8, 95% CI [49, 93.9]) compared to homologous vaccine recipients (GMT for BA1 23.3, 95% CI [16.7, 32.5] and for BA5 30.4, 95% CI [21.8, 42.5]; **Table 2**). Notably however, after three vaccine doses, neutralising activity against all the Omicron variants tested was markedly lower than the corresponding neutralising activity against the ancestral strain for all individuals (**Fig. 2B**; **Table 2**).

### Breakthrough infections induce higher nAb titres against the ancestral strain than against Omicron subvariants

A subset of our study cohort (n=61) acquired SARS-CoV-2 infection after enrolment. While genomic data related to the virus were not collected at the time of infection to identify the variant responsible for infection, the breakthrough infections coincided with the disappearance of the Delta variant and emergence and surge of Omicron variants in the community ^37,39^.

We measured binding antibody levels in plasma and nAb titres in sera collected 2-4 weeks after breakthrough infections in xvaccinated individuals who had previously recovered from infection with the ancestral variant (n=25) and previously SARS-CoV-2 naïve (n=26) individuals. Upon measuring IgG binding antibodies to RBD derived from different variants (ancestral, BA1, BA2), we found that post-infection RBD-binding levels for BA1 and BA2 were significantly increased compared to the corresponding pre-infection sera in the naïve participants who received either mRNA or AdVV primary vaccination (**Fig. 3A**; **Table 3**). For convalescent participants irrespective of primary vaccination the increase in binding IgG antibody levels to BA1 or BA2 RBD were not significant in individuals with breakthrough infections. Except for a few individuals, there was no significant increase in Ancestral RBD binding levels either after a breakthrough infection.

**Figure 3.**
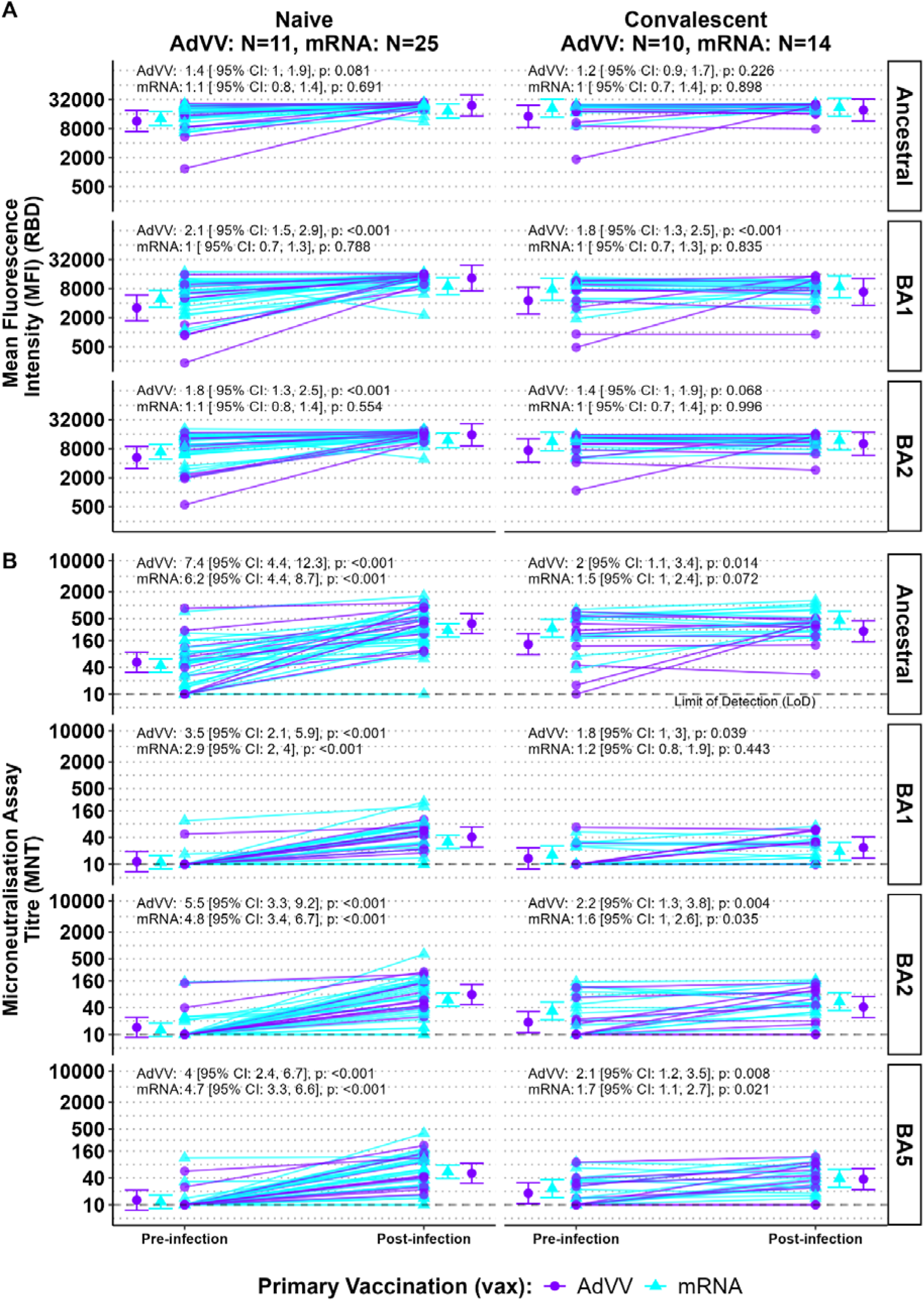
Differential boost of anti-RBD IgG binding antibodies and neutralising antibody titres after breakthrough infection in naïve and convalescent individuals. Anti-RBD IgG binding antibodies (**A**) to the ancestral strain and Omicron sub-variants (BA1 and BA2) and neutralising antibody titres (**B**) to the ancestral strain and Omicron sub-variants (BA1, BA2 and BA5) at pre-breakthrough infection (pre-infection) and post-breakthrough infection (post-infection) time points. Individuals are grouped based on their prior infection status (naïve and convalescent). Results are shown separately for each primary vaccination type (purple for AdVV and aqua for mRNA). The geometric mean and 95% confidence interval for each time point, within each sub-group are shown. The fold-changes from pre-infection to post-infection are shown for each primary vaccine type above each individual plot.

**Table 3.** Effect of breakthrough infections on levels of antigen-specific IgG binding antibody to RBD from SARS-CoV-2 Ancestral, BA1 and BA2 variants.

| SARS-CoV-2 strain | Original infection status | Primary vaccination | N | MFI<sup>†</sup><br>(Geometric mean [95% CI]) |  | Fold change in MFI pre- to post-infection (GMT [95% CI]) |
| --- | --- | --- | --- | --- | --- | --- |
|  |  |  |  | Pre-infection | Post-infection |  |
| Ancestral | Naive | AdVV | 11 | 11413<br>[6908, 18855] | 24066<br>[14567, 39759] | 2.1<br>[1.5, 3.0], p <0.001<sup>††</sup> |
| Ancestral | Convalescent | AdVV | 10 | 14302<br>[8447, 24215] | 19376<br>[11444, 32806] | 1.4<br>[0.9, 1.9], p=0.096 |
| Ancestral | Naive | mRNA | 25 | 12967<br>[9294, 18091] | 18476<br>[13243, 25778] | 1.4<br>[1.1, 1.8], p<0.002 |
| Ancestral | Convalescent | mRNA | 15 | 21131<br>[13747, 32481] | 22137<br>[14402, 34028] | 1.0<br>[0.8, 1.4], p=0.755 |
| BA1 | Naive | AdVV | 11 | 3183<br>[1721, 5886] | 13180<br>[7127, 24373] | 4.1<br>[2.4, 7.1], p<0.001 |
| BA1 | Convalescent | AdVV | 10 | 4532<br>[2378, 8636] | 6835<br>[3587, 13026] | 1.5<br>[0.9, 2.7], p=0.153 |
| BA1 | Naive | mRNA | 25 | 4902<br>[3260, 7370] | 8998<br>[5985, 13529] | 1.8<br>[1.3, 2.6], p<0.001 |
| BA1 | Convalescent | mRNA | 15 | 7811<br>[4614, 13224] | 8714<br>[5147, 14752] | 1.1<br>[0.7, 1.8], p=0.641 |
| BA2 | Naive | Ad VV | 11 | 5256<br>[3105, 8897] | 15598<br>[9215, 26401] | 3.0<br>[1.9, 4.5], p<0.001 |
| BA2 | Convalescent | AdVV | 10 | 7409<br>[4266, 12867] | 10154<br>[5847, 17634] | 1.4<br>[0.9, 2.1], p=0.162 |
| BA2 | Naive | mRNA | 25 | 6878<br>[4851, 9751] | 11900<br>[8393, 16871] | 1.7<br>[1.3, 2.3], p<0.001 |
| BA2 | Convalescent | mRNA | 15 | 11264<br>[7177, 17677] | 11813<br>[7527, 18538] | 1.0<br>[0.7, 1.5], p=0.796 |

<sup>†</sup>MFI: Median fluorescence intensity; CI: Confidence interval
<sup>††</sup>Using a regression model (detailed in the Statistical analysis section), p-value for difference from 1.0, signifying no change, in MFI values

In contrast to binding antibodies, nAb titres against all variants (ancestral, BA1, BA2 and BA5) were boosted after infection (post-infection) in all previously naïve individuals with a few exceptions (**Fig. 3B**; **Table 4**). In convalescent participants the changes in nAb titres for the variants tested varied, exhibiting an increase in some and a decrease in others after infection. Additionally, pre-infection nAb titres against the ancestral strain in convalescent AdVV recipients (GMT 132, 95% CI [77, 228]), and mRNA recipients (GMT 303, 95% CI [191, 481]) were on average higher than corresponding titres in naïve individuals (AdVV recipients: GMT 52, 95% CI [31, 88], and mRNA recipients: GMT 44, 95% CI [31, 62]). However, despite having breakthrough infections with an Omicron subvariant, there was a boost in nAb titres against the ancestral strain. This was particularly evident in the previously naïve group (**Fig. 3B**; **Table 4**).

**Table 4.** Effect of breakthrough infections on neutralising antibody titres to SARS-CoV-2 Ancestral, BA1, BA2 and BA5 variants.

| Target virus | Original infection status | Vaccination Type | N | Pre-infection GMT [95% CI]<sup>†</sup> | Post-infection GMT [95% CI] | Fold change pre- to post-infection [95% CI] |
| --- | --- | --- | --- | --- | --- | --- |
| Ancestral | Convalescent | AdVV | 10 | 132.4<br>[76.7, 228.4] | 259.6<br>[150.5, 448] | 2.0<br>[1.1, 3.4], p=0.014<sup>††</sup> |
| Ancestral | Naive | AdVV | 11 | 52.3<br>[31.1, 87.9] | 386.6<br>[229.8, 650.2] | 7.4<br>[4.4, 12.3], p<0.001 |
| Ancestral | Convalescent | mRNA | 14 | 303.3<br>[191.3, 480.9] | 460<br>[290.1, 729.3] | 1.5<br>[1.0, 2.4], p=0.072 |
| Ancestral | Naive | mRNA | 25 | 44.1<br>[31.2, 62.2] | 272.2<br>[192.8, 384.4] | 6.2<br>[4.4, 8.7], p<0.001 |
| BA1 | Convalescent | Ad VV | 10 | 13.5<br>[7.8, 23.3] | 23.8<br>[13.8, 41 .1] | 1.8<br>[1.0, 3.0], p=0.039 |
| BA1 | Naive | AdVV | 11 | 11 .5<br>[6.9, 1 9.4] | 40.9<br>[24.3, 68.8] | 3.5<br>[2.1, 5.9], p<0.001 |
| BA1 | Convalescent | mRNA | 14 | 16.4<br>[10.3, 26.0] | 19.6<br>[12.4, 31 .1] | 1.2<br>[0.8, 1.9], p=0.443 |
| BA1 | Naive | mRNA | 25 | 11 .2<br>[7.9, 15.8] | 32<br>[22.7, 45.2] | 2.9<br>[2.0, 4.0], p<0.001 |
| BA2 | Convalescent | AdVV | 10 | 1 8.9<br>[10.9, 32.6] | 41 .5<br>[24.1, 71.7] | 2.2<br>[1.3, 3.8], p=0.004 |
| BA2 | Naive | AdVV | 11 | 14.4<br>[8.6, 24.31] | 79.3<br>[47.1, 133.8] | 5.5<br>[3.3, 9.2] p<0.001 |
| BA2 | Convalescent | mRNA | 14 | 33.7<br>[21.3, 53.5] | 54.9<br>[34.6, 87.1] | 1.6<br>[1.0, 2.6], p=0.035 |
| BA2 | Naive | mRNA | 25 | 12.7<br>[9.0, 1 8] | 60.6<br>[42.9, 85.6] | 4.8<br>[3.4, 6.7], p<0.001 |
| BA5 | Convalescent | AdVV | 10 | 18.2<br>[10.5, 31 .41] | 37.6<br>[21.8, 64.9] | 2.1<br>[1.2, 3.5], p=0.008 |
| BA5 | Naive | AdVV | 11 | 12.7<br>[7.6, 21 .4] | 51 .2<br>[30.4, 86.1] | 4<br>[2.4, 6.7], p<0.001 |
| BA5 | Convalescent | mRNA | 14 | 23.1<br>[14.5, 36.6] | 39.4<br>[24.8, 62.4] | 1.7<br>[1.1, 2.7], p=0.021 |
| BA5 | Naive | mRNA | 25 | 11.7<br>[8.3, 16.5] | 55.0<br>[39, 77.6] | 4.7<br>[3.3, 6.6], p<0.001 |

<sup>†</sup>GMT: Geometric mean neutralising titres; CI: confidence interval
<sup>††</sup>Using a regression model (detailed in the Statistical analysis section), p-value for difference from 1.0, signifying no change, in GMT values

Stratification of both the naïve and convalescent individuals based on receiving homologous or heterologous vaccination showed that average fold-increase in nAb titres from pre-infection to post-infection timepoints did not differ between the two vaccine groups (**Table 4**). As noted above, infection occurring during high levels of Omicron transmission in the community was associated with a boosting of nAb titres against both the ancestral virus and Omicron subvariants in the naïve participants (**Fig. 3B**; **Table 4**). These observations are consistent with enhanced response to the original vaccine antigen suggesting that antigen imprinting by vaccination with the ancestral strain had occurred.

## Discussion

In this study we used sera/plasma samples collected from individuals living in Australia, a low SARS-CoV-2 transmission country in the two first years of the COVID-19 pandemic, to perform comprehensive analysis of binding and neutralising antibody levels in response to COVID-19 vaccines and breakthrough infections. The two salient findings of our study were, first, the need of a booster (third dose) of vaccine for the generation of neutralising activity against Omicron variants, and second, the dominant boosting of neutralising activity against the ancestral strain following infection with Omicron variants, indicative of imprinting of the immune response to the original antigen. If imprinting does occur in the context of vaccines, studies investigating the antigenic “distance” required to circumvent the imprinting would be of great interest for the design of future vaccines against SARS-CoV-2.

Several studies of vaccine responses to two-dose vaccination have been reported. However, most have been conducted in the setting of high (or unknown) levels of viral infections in the community or of work-related exposure in healthcare workers. This factor can influence immune responses to viral antigens. Here, we confirm previous findings of a robust rise in levels of binding antibodies to ancestral Spike antigens after two vaccine doses^33,40,41^ in a population with low levels of community transmission. We also found that in the absence of exposure to the virus in a region with absent or extremely low levels of community transmission, the antibody responses induced by vaccination differed between the infection-naive and COVID-19 convalescent individuals as has been reported in conditions of ambiguous transmission^41,42^.

Previous studies have shown that a third booster dose increases nAb titres and binding antibodies above the levels achieved by two doses of vaccine^33,43^. Amongst the infection-naive individuals, the third vaccine dose boosted neutralising activity against the variant virus strains as well as the ancestral strain. However, the third vaccine dose did not increase binding antibody levels in this group. The discordance between the binding antibody and nAb response to the vaccine likely reflects the broad targeting of the former to a range of Spike protein antigens, compared to the narrower subset of neutralising Ab targets. Furthermore, in convalescent individuals the booster dose produced minimal change in binding and nAb titres overall, presumably due to the higher absolute neutralising levels already achieved by hybrid immunity in convalescent individuals who were subsequently vaccinated. Since the primary vaccinations in our study population were consistently either two doses of AdVV or two doses of mRNA vaccine, followed by an mRNA booster as the third dose in all participants, we were able to compare antibody responses after heterologous (AdVV/AdVV/mRNA) versus homologous (mRNA/mRNA/mRNA) vaccination. Previous reports demonstrate that heterologous vaccination induces broader and more durable antibody responses^44,45^. Of note in the present study, in infection-naïve individuals, a third vaccine dose was required to generate neutralising activity against Omicron subvariants, irrespective of primary vaccination type. Our data show that in naïve individuals, heterologous vaccination induced a greater boost in neutralising antibody levels to the ancestral strain of SARS- CoV-2 than was observed after homologous vaccination. In addition, when we stratified the convalescent cohort by vaccine received (AdVV or mRNA), there was a significant boost of neutralising activity to both the ancestral virus and variant strains in the AdVV recipients after the third vaccine dose (mRNA, heterologous vaccination) which supports previous reports that heterologous vaccination has the capacity to induce better cross-variant neutralisation^46^.

In individuals previously vaccinated and boosted with antigens derived from the ancestral strain of SARS-CoV-2, breakthrough infections with variants stimulate a *de novo* expansion of B cells targeting the altered viral spike glycoprotein but, at the same time, also an expansion of cross-reactive B and T cells previously sensitised to shared epitopes^21,47,48^. “Imprinting” of immune responses refers to the concept whereby following first exposure to an antigen, immune responses to subsequent exposure to a closely related new antigen predominantly targets epitopes that are shared with the original antigen. Evidence for immunological imprinting has been found with Omicron infections^11,12,21,49–52^. In these studies, in individuals previously vaccinated with the spike protein from the ancestral strain, Omicron infections were associated with a boost in neutralising Ab titres against the ancestral strain as well as the infecting Omicron strain^12,21,23^. Our data are consistent with these reports. We demonstrated that Omicron infections boosted nAb titres against the ancestral strain as well as, or better than, nAb titres against the infecting Omicron strains (**Fig. 3B**).

The mechanisms underlying the phenomenon of imprinting await conclusive explanation, however, it has been proposed that epitope masking and feedback inhibition by pre-existing Abs may impede the recruitment of naive B cells specific to novel epitopes on variant spike proteins^23,53,54^. Interestingly, in a study reported by Yisimayi and colleagues^55^, robust variant-specific responses were seen after Omicron infections in individuals who previously received inactivated SARS-CoV-2 vaccine, suggesting that inactivated virus vaccines may leave fainter immunological imprints compared with mRNA or vectored vaccines.

The clinical significance of immunological imprinting is as yet uncertain. Booster vaccines containing spike proteins from BA.5 and XBB.1.5 remain very effective in preventing severe disease and deaths caused by these variants^56–62^, suggesting that Abs directed against shared epitopes with the strain that imprints the immune response contribute to protection provided by the variant booster vaccines against severe outcomes.

Our study, that leveraged access to an increasingly rare COVID19-naïve population has some limitations. While the findings of the study give a unique longitudinal perspective of antibody responses following primary and booster vaccination and after breakthrough infection, the sample size is relatively small. This reflects the limitations of conducting research in a rapidly changing environment as a result of the pandemic as well as specific local factors, such as repeated lockdowns that prevented travel and visits to the clinic. As a result, we could not explore potential differences in antibody titres between males and females. The study did not include the elderly or children. This may limit the scope and generalisability of our results, but the consistency of the responses within each subgroup supports internal validity of the data and lends strength to our conclusions. Because the reporting of SARS-CoV-2 infections in Australia changed from PCR in centralised laboratories to self- testing with Rapid Antigen Tests (RATs), we do not have definitive information on the infective variant of the breakthrough infections. However, based on transmission data in the Australia and locally in Melbourne, the timing of breakthrough infections that occurred in this cohort coincides with epidemiological data where 99% of the infections were caused by Omicron variants. B cell responses were not characterised in this study because the rapid implementation of research studies in the early phases of the SARS- CoV-2 pandemic did not allow us time to establish the necessary protocols. However, analysis of B cell proliferation in a subset of our participants using a mathematical model of in-host immune cell kinetics estimated that mRNA vaccines induced 2.1 times higher memory B cell proliferation than AdVV vaccines after adjusting for age, interval between doses and priming dose. Additionally, extending the duration between the priming dose and second vaccine dose beyond 28 days boosted neutralising antibody production per plasmablast concentration by 30%^63^. Additional analyses could have provided validation of our data and potentially elucidated underlying mechanisms for the observed patterns of immune responses to the vaccines and infections.

In conclusion, our study of vaccine-induced immune responses is unique because it was conducted in COVID-19-naïve and post-COVID-19 infected individuals in a setting where the confounding effects of community transmission and unintentional exposure to SARS-CoV-2 infections was circumvented. Thus, we characterised *de novo* antibody responses to three doses of vaccines as well as responses to infection with SARS-CoV-2 variants. We have demonstrated that, in these conditions, two doses of vaccines were insufficient for generation of nAb responses to the variant viruses, that a 3^rd^ dose of either a heterologous or homologous vaccine induced equivalent neutralising antibody responses in both infection-naïve and convalescent individuals and, that infection of vaccinated individuals with SARS-CoV-2 boosts levels of nAbs against the infecting variant in addition to the original vaccine virus, indicative of immune imprinting. Immune imprinting needs to be addressed in vaccine design and vaccination programs because the first experience with SARS-CoV-2 in different populations around the world varies greatly, as does the context and nature of subsequent exposure to the virus.

## Acknowledgements

We wish to thank Jason Tye-Din, Anne Hart, Maureen Ford, Siavash Foroughi, Shazia Ruybal-Pesantes, all members of the COVID PROFILE consortium, the study coordinators (Chioma Uzoho, Fae Brigino and Senila Gunawardane) and all the study participants in the COVID Profile and DISCOVER-HCP studies. Study data were collected and managed using REDCap electronic data capture tools hosted at the University of Melbourne. We thank the team for their assistance in creation and management of the database.

## Author contributions

EE, SM, KS, VB, IM conceived and conducted the COVID Profile and DISCOVER-HCP studies. NW, RM, HD, LYYL, LR assisted with cohort studies, EE, SM, FM, LYYL, RM, NW and EC analysed samples and generated the data. EE, AM, PE, SB, and DP performed the data and statistical analyses. EE, SM, KS and PE wrote the first draft of the manuscript. KS, VB, IM, and DP provided guidance and helpful discussion and edited the manuscript. We acknowledge invaluable assistance provided by Barbara Scher (Department of Infectious Diseases, The Peter Doherty Institute for Infection and Immunity) in the establishment, conduct and governance of the DISCOVER-HCP study.

## Data availability

All data produced in the present study are available upon reasonable request to the authors

