## Supplemental files for "Antibody responses following COVID-19 vaccination and breakthrough infections in naïve and convalescent individuals suggests imprinting to the ancestral strain of SARS-CoV-2"

**Mahanty *et al.***

**Supplementary Tables and Figures**

**Tables S1 – S5**

**Figures S1 – S6 with legends**

**Supplementary Table S1a:** All participants (N=126) grouped by prior study infection status and primary vaccine time

|  | **Naïve^¶^** | | | **Convalescent^§^** | | |
| --- | --- | --- | --- | --- | --- | --- |
|  | **AdVV* (N=28)** | **mRNA** (N=44)** | **Total (N=72)** | **AstraZeneca**  **(AdVV) (N=25)** | **Pfizer**  **(mRNA) (N=29)** 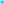 | **Total (N=54)** |
| **Gender** 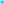 | | | | | | |
| Female | 19 (68%) | 30 (68%) | 49 (68%) | 10 (40%) | 21 (72%) | 31 (57%) |
| Male | 9 (32%) | 14 (32%) | 23 (32%) | 15 (60%) | 8 (28%) | 23 (43%) |
| **Age (years)** | | | | | | |
| Median (IQR) | 52.0 (37.8 - 59.3) | 39.0 (31.0 - 49.0) | 44.5 (32.0 - 53.3) | 59.0 (53.0 - 61.0) | 46.0 (36.0 - 53.0) | 52.5 (39.5 - 59.0) |
| **COVID-19 Vaccination (booster)** | | | | | | |
| Pfizer | 20 (71%) | 36 (82%) | 56 (78%) | 20 (80%) | 18 (62%) | 38 (70%) |
| Moderna | 5 (18%) | 4 (9%) | 9 (12%) | 5 (20%) | 7 (24%) | 12 (22%) |
| Not Vaccinated | 2 (7%) | 0 (0%) | 2 (3%) | 0 (0%) | 4 (14%) | 4 (7%) |
| Missing | 1 (3.6%) | 4 (9.1%) | 5 (6.9%) | 0 (0%) | 0 (0%) | 0 (0%) |
| **Baseline visit** | | | | | | |
| yes | 20 (71%) | 31 (70%) | 51 (71%) | 24 (96%) | 23 (79%) | 47 (87%) |
| Missing | 8 (28.6%) | 13 (29.5%) | 21 (29.2%) | 1 (4.0%) | 6 (20.7%) | 7 (13.0%) |
| **Post 2nd vaccine visit** | | | | | | |
| yes | 21 (75%) | 33 (75%) | 54 (75%) | 24 (96%) | 23 (79%) | 47 (87%) |
| Missing | 7 (25.0%) | 11 (25.0%) | 18 (25.0%) | 1 (4.0%) | 6 (20.7%) | 7 (13.0%) |
| **Pre-booster vaccine visit** | | | | | | |
| Yes | 20 (71%) | 32 (73%) | 52 (72%) | 20 (80%) | 19 (66%) | 39 (72%) |
| Missing | 8 (28.6%) | 12 (27.3%) | 20 (27.8%) | 5 (20.0%) | 10 (34.5%) | 15 (27.8%) |
| **Post-booster vaccine visit** | | | | | | |
| yes | 20 (71%) | 32 (73%) | 52 (72%) | 20 (80%) | 19 (66%) | 39 (72%) |
| Missing | 8 (28.6%) | 12 (27.3%) | 20 (27.8%) | 5 (20.0%) | 10 (34.5%) | 15 (27.8%) |
| **Pre-breakthrough infection visit** | | | | | | |
| yes | 11 (39%) | 25 (57%) | 36 (50%) | 10 (40%) | 15 (52%) | 25 (46%) |
| Missing | 17 (60.7%) | 19 (43.2%) | 36 (50.0%) | 15 (60.0%) | 14 (48.3%) | 29 (53.7%) |
| **Post-breakthrough infection visit** | | | | | | |
| yes | 11 (39%) | 25 (57%) | 36 (50%) | 10 (40%) | 15 (52%) | 25 (46%) |
| Missing | 17 (60.7%) | 19 (43.2%) | 36 (50.0%) | 15 (60.0%) | 14 (48.3%) | 29 (53.7%) |
| ^¶^No previous SARA-CoV-2 infections; ^§^Dcoumented previous infection; * ChAdOX1 (AstraZeneca), 2 doses followed by BNT162b2 (mRNA; Pfizer); ** BNT162b2 (mRNA, Pfizer) 3 doses | | | | | | |

**Supplementary Table 1b:** Demographic and vaccine history of 61 participants who had documented breakthrough infections with SARS-CoV-2

|  | **Naive** | | | **Convalescent** | | |
| --- | --- | --- | --- | --- | --- | --- |
|  | **AdVV (N=11)** | **mRNA) (N=25)** | **Total (N=36)** | **AdVV (N=10)** | **mRNA (N=15)** | **Total (N=25)** |
| **Gender** | | | | | | |
| Female | 7 (64%) | 18 (72%) | 25 (69%) | 5 (50%) | 11 (73%) | 16 (64%) |
| Male | 4 (36%) | 7 (28%) | 11 (31%) | 5 (50%) | 4 (27%) | 9 (36%) |
| **Age (years)** | | | | | | |
| Median (IQR) | 53.0 (43.0 - 63.0) | 43.0 (31.0 - 49.0) | 45.0 (31.8 - 52.3) | 59.5 (56.8 - 61.8) | 49.0 (29.5 - 55.0) | 55.0 (42.0 - 59.0) |
| **COVID-19 Vaccination (booster)** | | | | | | |
| Pfizer | 5 (45%) | 19 (76%) | 24 (67%) | 5 (50%) | 9 (60%) | 14 (56%) |
| Moderna | 4 (36%) | 4 (16%) | 8 (22%) | 5 (50%) | 6 (40%) | 11 (44%) |
| Not Vaccinated | 2 (18%) | 0 (0%) | 2 (6%) | 0 (0%) | 0 (0%) | 0 (0%) |
| Missing | 0 (0%) | 2 (8.0%) | 2 (5.6%) | 0 (0%) | 0 (0%) | 0 (0%) |
| **Baseline visit** | | | | | | |
| yes | 3 (27%) | 15 (60%) | 18 (50%) | 9 (90%) | 9 (60%) | 18 (72%) |
| Missing | 8 (72.7%) | 10 (40.0%) | 18 (50.0%) | 1 (10.0%) | 6 (40.0%) | 7 (28.0%) |
| **Post 2nd vaccine visit** | | | | | | |
| yes | 4 (36%) | 15 (60%) | 19 (53%) | 9 (90%) | 9 (60%) | 18 (72%) |
| Missing | 7 (63.6%) | 10 (40.0%) | 17 (47.2%) | 1 (10.0%) | 6 (40.0%) | 7 (28.0%) |
| **Pre-booster vaccine visit** | | | | | | |
| yes | 4 (36%) | 15 (60%) | 19 (53%) | 6 (60%) | 9 (60%) | 15 (60%) |
| Missing | 7 (63.6%) | 10 (40.0%) | 17 (47.2%) | 4 (40.0%) | 6 (40.0%) | 10 (40.0%) |
| **Post-booster vaccine visit** | | | | | | |
| yes | 4 (36%) | 15 (60%) | 19 (53%) | 6 (60%) | 9 (60%) | 15 (60%) |
| Missing | 7 (63.6%) | 10 (40.0%) | 17 (47.2%) | 4 (40.0%) | 6 (40.0%) | 10 (40.0%) |
| **Pre-breakthrough infection visit** | | | | | | |
| yes | 11 (100%) | 25 (100%) | 36 (100%) | 10 (100%) | 15 (100%) | 25 (100%) |
| **Post-breakthrough infection visit** | | | | | | |
| yes | 11 (100%) | 25 (100%) | 36 (100%) | 10 (100%) | 15 (100%) | 25 (100%) |

**Supp table S2a**. Antibody responses to SARS-CoV-2 RBD protein following 2 doses of vaccines, by prior infection-status and vaccine type

| **Antibody type** | **Original infection status** | **Vaccination Type** | **N** | **Geometric Mean of MFI^¶^**  **[95% Confidence Interval (CI)]** | | **Fold change [95% CI]** |
| --- | --- | --- | --- | --- | --- | --- |
|  |  |  |  | **Pre-Vax** | **Post-2nd vaccine** | **Post-2nd vaccine / Pre-Vax** |
| IgG | Naive | AdVV | 20 | 176.1 [118.7, 261.2] | 11154.8 [7519.6, 16547.5] | 63.3 [38.5, 104.3], p: <0.001 |
| IgG | Naive | mRNA | 31 | 111.1 [81, 152.6] | 16704.5 [12169.3, 22929.8] | 150.3 [100.7, 224.3], p: <0.001 |
| IgG | Convalescent | AdVV | 24 | 3388.3 [2363.9, 4856.5] | 17987.8 [12549.6, 25782.4] | 5.3 [3.4, 8.4], p: <0.001 |
| IgG | Convalescent | mRNA | 23 | 3426.1 [2371.9, 4948.9] | 23603.4 [16340.5, 34094.5] | 6.9 [4.3, 11], p: <0.001 |
| IgM | Naive | AdVV | 20 | 512.5 [348.1, 754.4] | 465.6 [316.3, 685.5] | 0.9 [0.6, 1.4], p: 0.652 |
| IgM | Naive | mRNA | 31 | 487.1 [357.1, 664.6] | 567.9 [416.3, 774.8] | 1.2 [0.8, 1.6], p: 0.369 |
| IgM | Convalescent | AdVV | 24 | 305.1 [214.3, 434.2] | 741.6 [521, 1055.5] | 2.4 [1.7, 3.6], p: <0.001 |
| IgM | Convalescent | mRNA | 23 | 328 [228.7, 470.4] | 635.3 [443, 911.1] | 1.9 [1.3, 2.9], p: <0.001 |
| IgA | Naive | AdVV | 20 | 337.6 [222.3, 512.6] | 553.6 [364.6, 840.6] | 1.6 [1.1, 2.4], p: 0.012 |
| IgA | Naive | mRNA | 31 | 216 [154.4, 302.1] | 1335.1 [954.6, 1867.2] | 6.2 [4.5, 8.4], p: <0.001 |
| IgA | Convalescent | AdVV | 24 | 1599.7 [1092.6, 2342.2] | 2313.9 [1580.4, 3387.8] | 1.4 [1, 2.1], p: 0.04 |
| IgA | Convalescent | mRNA | 23 | 1471 [996.5, 2171.5] | 3467.6 [2349.1, 5118.8] | 2.4 [1.6, 3.4], p: <0.001 |
| ^¶^Median fluorescence intensity in a multiplex bead array assay (See Methods) | | | | | | |

**Supplementary Table S2b**. Comparison of IgG to SARS-CoV-2 RBD protein between infection-naïve and previously infected recipients by vaccine type

| **Antibody type** | **Vaccination Type** | **Post-2nd vaccine geometric mean**  **[95% Confidence Interval (CI)]** | | **Fold change [95% CI]** |
| --- | --- | --- | --- | --- |
|  |  | **Naive** | **Convalescent** | **Convalescent / Naive** |
| IgG | AdVV | 11154.8 [7519.6, 16547.5] | 17987.8 [12549.6, 25782.4] | 1.6 [0.9, 2.7] |
| IgG | mRNA | 16704.5 [12169.3, 22929.8] | 23603.4 [16340.5, 34094.5] | 1.4 [0.9, 2.3] |

**Supplementary Table S3**. Comparison of neutralising titres to Ancestral strain of SARS-CoV-2 in infection-naïve and previously infected recipients by vaccine type

| **Original infection status** | **Vaccination Type** | **N** | **Geometric Mean [95% Confidence Interval (CI)]** | | **Fold change pre-post [95% CI]** | **Fold change ratio Convalescent:**  **Naive [95% CI]** | **Post-2nd vaccine GM Ratio mRNA:AdVV [95% CI]** |
| --- | --- | --- | --- | --- | --- | --- | --- |
|  |  |  | **Pre-Vax** | **Post-2nd vaccination** |  |  |  |
| Naive | AdVV | 19 | 10 [LoD] | 46.9 [33.5, 65.7] | 4.7 [3.4, 6.6], p: <0.001* | Reference | Reference^ |
| Convalescent | AdVV | 24 | 29.8 [22.1, 40.2] | 519.7 [385.1, 701.3] | 17.5 [13, 23.5], p: <0.001 | 3.7 [2.4, 5.8] | Reference^^ |
| Naive | mRNA | 31 | 10 [LoD] | 100.9 [77.5, 131.3] | 10.1 [7.8, 13.1], p: <0.001* | Reference | 2.1 [1.4, 3.3]^ |
| Convalescent | mRNA | 23 | 22 [16.2, 29.9] | 1496.4  [1101.7, 2032.6] | 68.1 [50.2, 92.2], p: <0.001 | 6.7 [4.5, 10.1] | 2.9 [1.9, 4.4]^^ |

**Supplementary Table S4**. Comparison of IgG to SARS-CoV-2 RBD protein between infection-naïve and previously infected recipients by vaccine type

| **Original infection status** | **Vaccination Type** | **N** | **Geometric Mean [95% Confidence Interval (CI)]** | | **Fold change pre-post [95% CI]** | **Fold change ratio Convalescent:**  **Naive [95% CI]** | **Post-2nd vaccine GM Ratio mRNA:AdVV [95% CI]** |
| --- | --- | --- | --- | --- | --- | --- | --- |
|  |  |  | **Pre-Vax** | **Post-2nd vaccination** |  |  |  |
| Naive | AdVV | 20 | 60.7  [41.6, 88.6] | 4324.4  [2964.5, 6308] | 71.2 [44.7, 113.5], p: <0.001 | Reference | Reference^ |
| Convalescent | AdVV | 24 | 584.3  [413.9, 824.7] | 9076.8  [6430.6, 12811.9] | 15.5 [10.2, 23.8], p: <0.001 | 4.6 [2.4, 8.6] | Reference^^ |
| Naive | mRNA | 31 | 73.7  [54.4, 99.8] | 8100  [5981.1, 10969.5] | 109.9 [75.6, 159.8], p: <0.001 | Reference | 1.9 [1.1, 3.1]^ |
| Convalescent | mRNA | 23 | 635.3  [446.8, 903.4] | 15090.6  [10612.2, 21458.8] | 23.8 [15.4, 36.7], p: <0.001 | 4.6 [2.6, 8.2] | 1.7 [1.0, 2.7]^^ |

**Supplementary Table 5**: Pre-vaccination (pre-vax) and post-2^nd^ vaccination neutralising antibody titres against the Delta variant (A) and post-2^nd^ vaccination comparison to the Ancestral Strain (B)

**Table 5A**

| **Original infection status** | **Vaccination Type** | **N** | **Geometric Mean [95% Confidence Interval (CI)]** | | **Fold change pre-post vaccination [95% CI]** | **Magnitude difference in fold change Convalescent:Naive [95% CI]** |
| --- | --- | --- | --- | --- | --- | --- |
|  |  |  | **Pre-Vax** | **Post-2nd vaccination** |  |  |
| Naive | AdVV | 19 | 10 [LoD] | 29 [20.7, 40.6] | 2.9 [2.1, 4], p: <0.001* | 4 [2.5, 6.2] p:<0.001 |
| Convalescent | AdVV | 24 | 20.3 [15, 27.4] | 234.1 [173.5, 315.9] | 11.5 [8.6, 15.5], p: <0.001 |  |
| Naive | mRNA | 31 | 10 [LoD] | 37.4 [28.7, 48.7] | 3.7 [2.9, 4.9], p: <0.001* | 11.4 [7.7, 17.1] p:<0.001 |
| Convalescent | mRNA | 23 | 12 [8.9, 16.3] | 514.4 [378.7, 698.7] | 42.8 [31.6, 58], p: <0.001 |  |

**Table 5B**

| **Original infection status** | **Vaccination Type** | **Geometric Mean [95% Confidence Interval (CI)]** | | **Fold change [95% CI]** |
| --- | --- | --- | --- | --- |
|  |  | **Ancestral Strain** | **Delta Strain** | **Ancestral/Delta** |
| Naive | AdVV | 46.9 [33.5, 65.7] | 29 [20.7, 40.6] | 1.6 [1.2, 2.3], p: 0.005 |
| Convalescent | AdVV | 519.7 [385.1, 701.3] | 234.1 [173.5, 315.9] | 2.2 [1.6, 3], p: <0.001 |
| Naive | mRNA | 100.9 [77.5, 131.3] | 37.4 [28.7, 48.7] | 2.7 [2.1, 3.5], p: <0.001 |
| Convalescent | mRNA | 1496.4 [1101.7, 2032.6] | 514.4 [378.7, 698.7] | 2.9 [2.1, 3.9], p: <0.001 |

**Supplementary Figure S1**

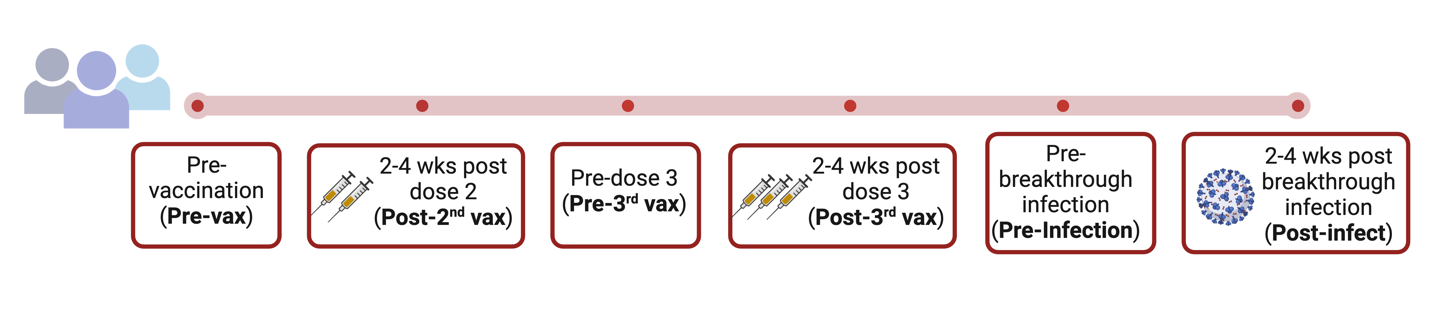

**Figure** **S1:** Summary of sample collection time points

**Supplementary Figure S2**

**Figure S2: S**chema of participant enrolment and selection of samples for IgG antibody analysis at selected study points

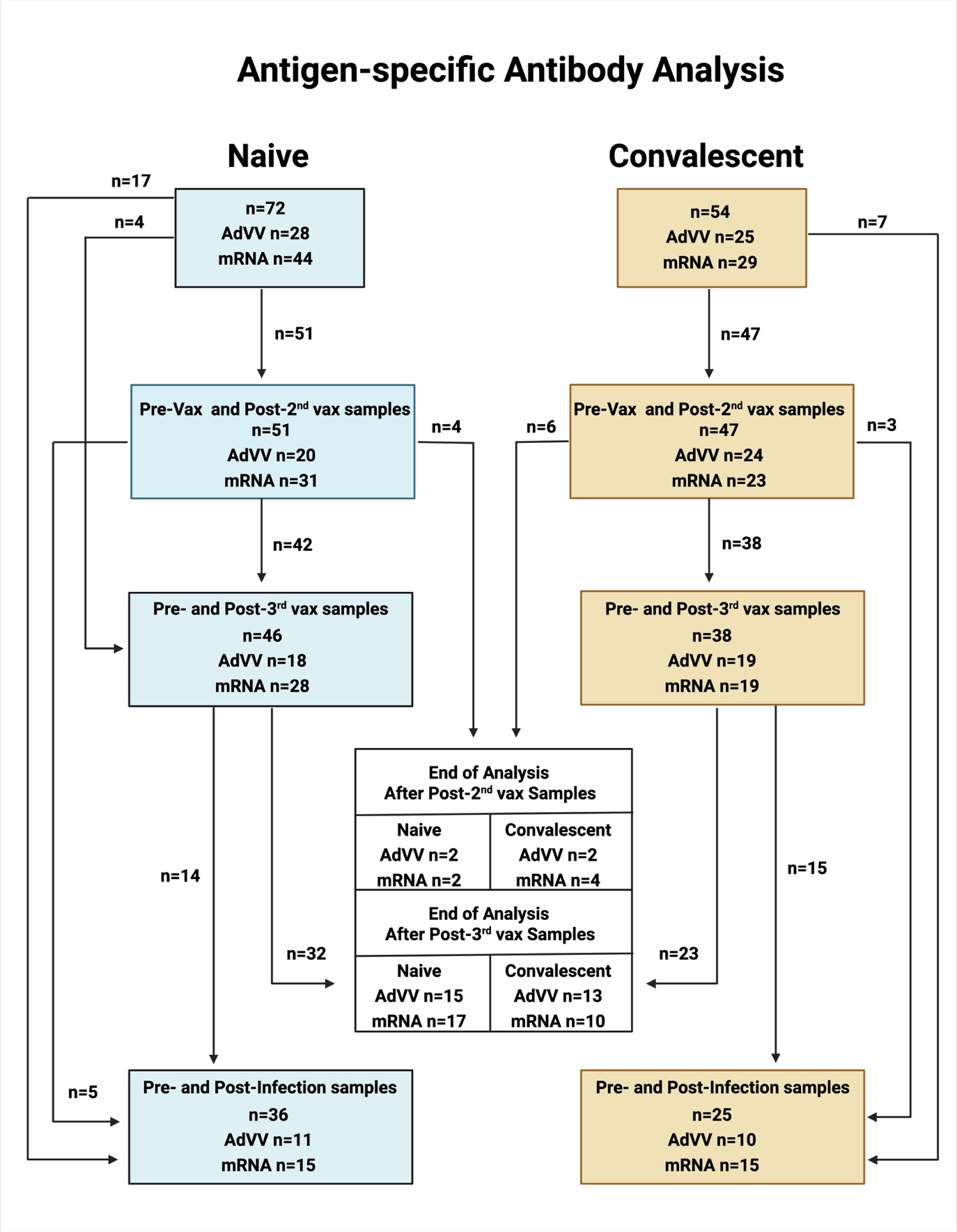

**Supplementary Figure S3**

**Figure S3:** Schema of participant enrolment and selection of samples for neutralising antibody analysis at selected study points

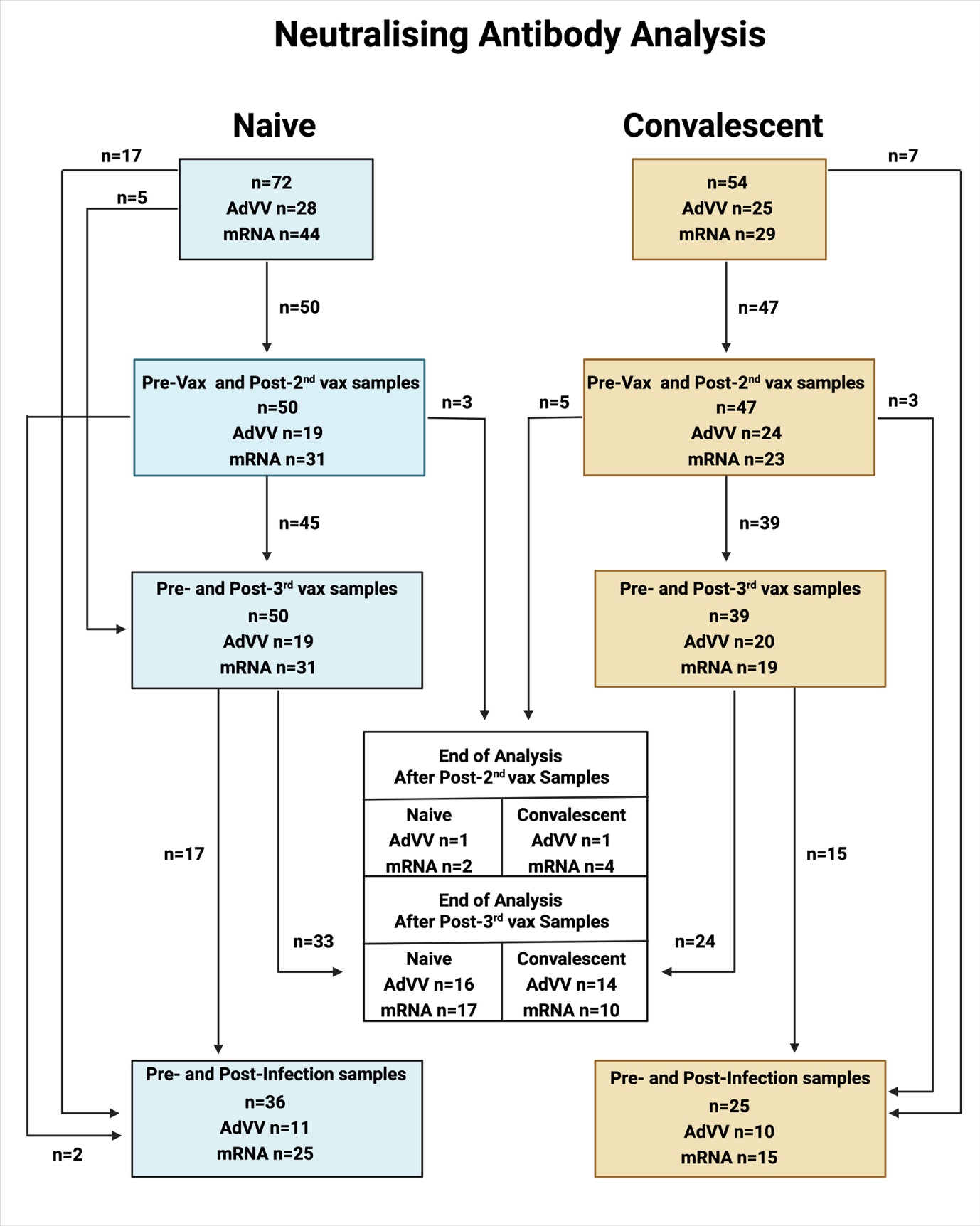

**Supplementary Figure S4**

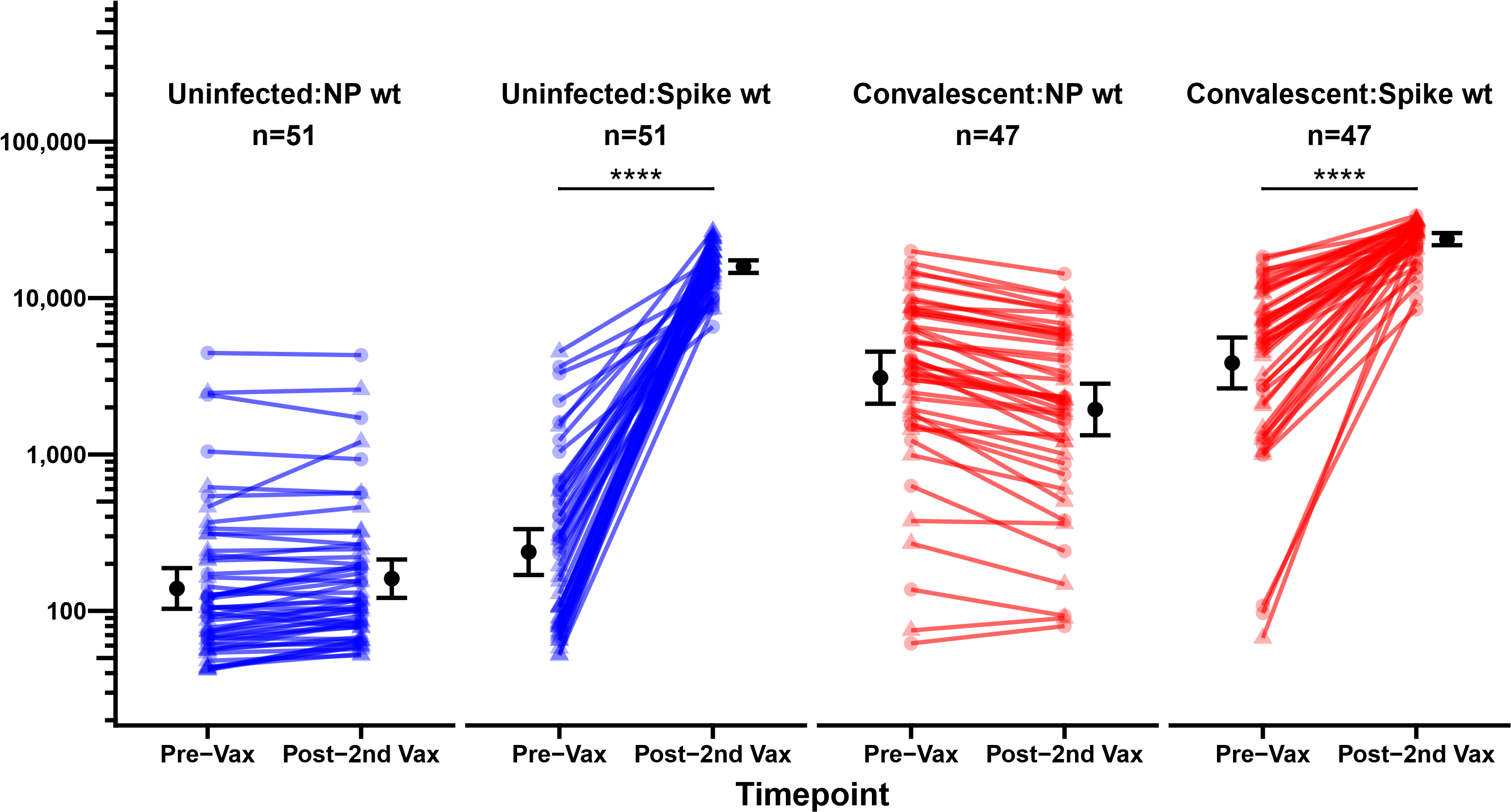

Median fluorescence intensity

)

(

MFI

**Figure S4. Anti-NP IgG binding antibodies to Ancestral proteins after two COVID-19 vaccine doses**. Anti-NP and anti-Spike IgG binding antibodies (MFi) at pre-vaccination and post-2^nd^ vaccination time points for each individual. Participants are stratified on vaccine type (filled triangles for AdVV and filled circles for mRNA) and prior infection status (blue for Naïve participants and red for Convalescent participants). Black circles with errorbars represent the geometric mean and 95% confidence intervals for the cohort. **** p<0.001 in paired two-sample Wilcoxon signed-rank tests.

**Supplementary Figure S5**

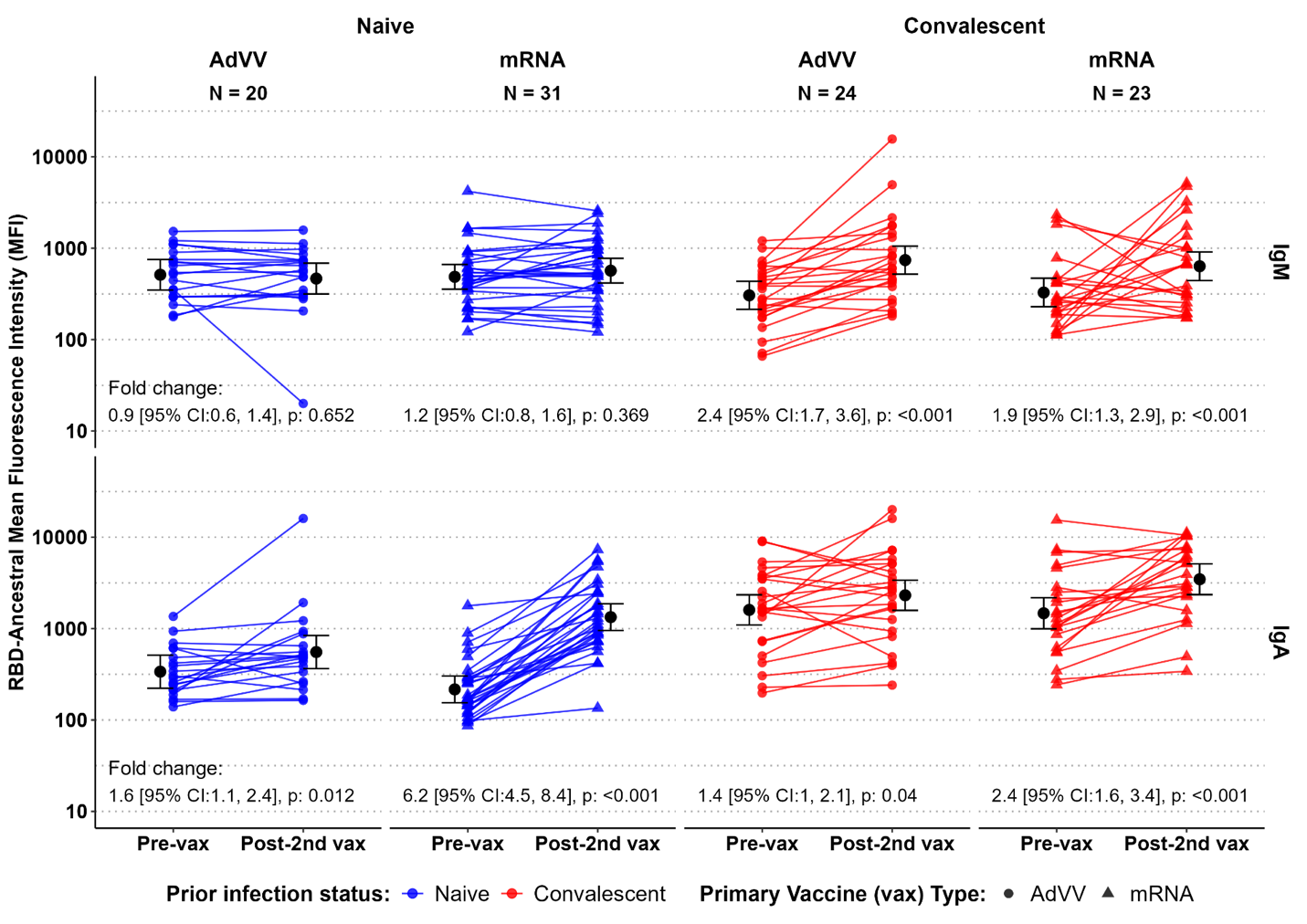

**Figure S5:** Anti-RBD IgM and IgA binding antibody MFI levels after two COVID-19 vaccine doses. Anti-RBD (ancestral) antibody MFI levels for IgM (top panel) and IgA (bottom panel) pre-vaccination (pre-vax) and post-2^nd^ vaccination (Post-2^nd^-vax) are shown for all participants (blue for Naïve participants and red for convalescent participants) with the geometric mean and 95% confidence interval for each time point shown in black.

**Supplementary Figure S6**

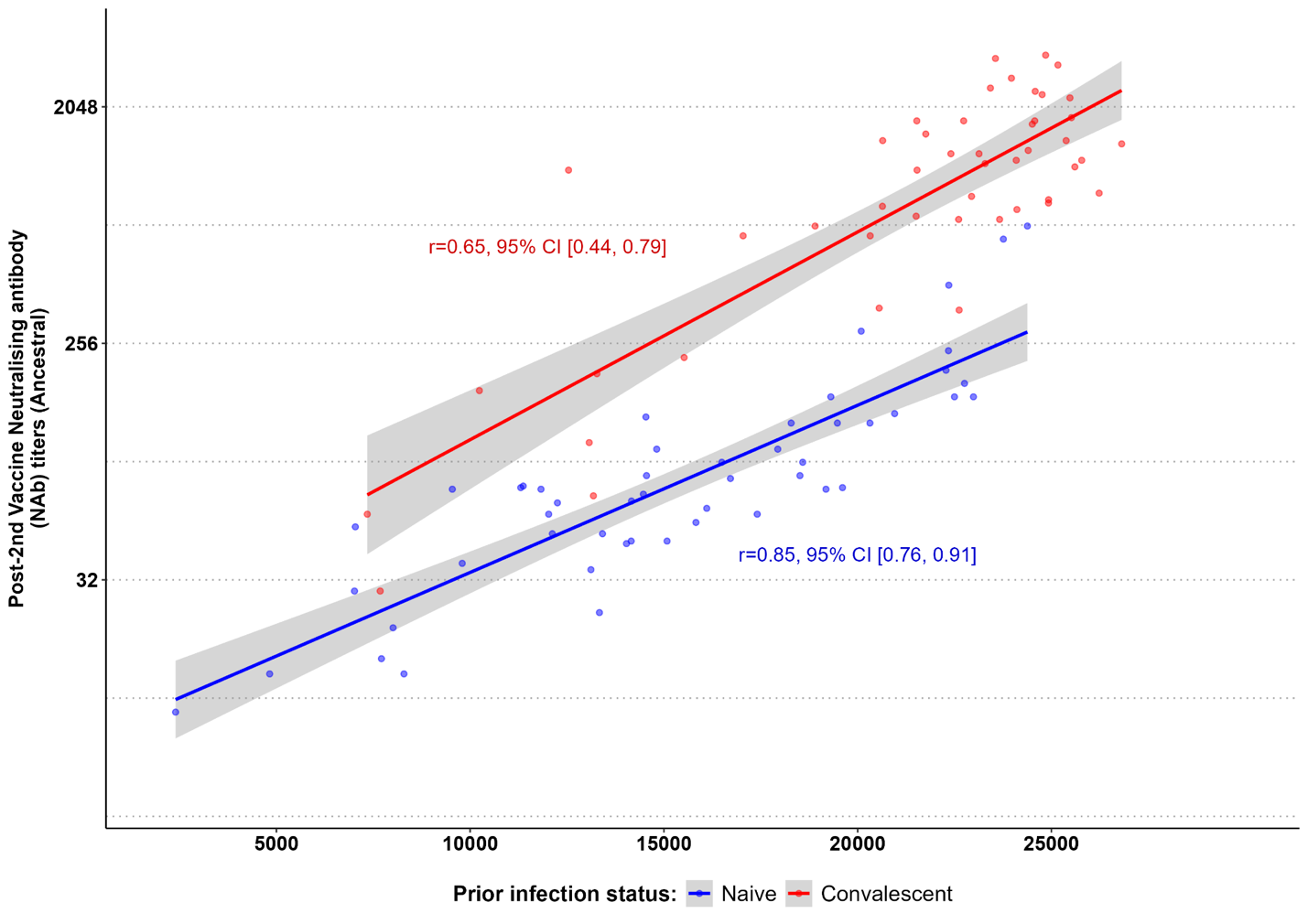

**Figure S6** Spearman correlation of post-2^nd^ vaccine anti-RBD (ancestral) IgG binding antibody MFI levels to neutralising (ancestral) antibody titres. Each individual is represented as a single dot (blue for Naïve participants and red for Convalescent participants).
